# Synergistic Effect of VSL#3^™^ and Vedolizumab for Ulcerative colitis: Preliminary Evidence from a Real-world Single-Arm study

**DOI:** 10.64898/2026.07.23.26358760

**Authors:** Qiao Yu, Jiakai Luo, Xiaoying Wang, Dingting Xu, Hanyun Zhang, Miaoyan Chen, Shuyan Li, Parnia Ghanad, Maryam Maleki Goli, Yan Chen

## Abstract

**Objective:** Vedolizumab (VDZ) is effective in ulcerative colitis (UC), but its onset of action may be relatively slow during induction therapy. VSL#3™, a high-potency multi-strain probiotic, may provide synergistic effects through microbiota modulation and immune regulation. This preliminary real-world study aimed to evaluate the efficacy and safety of VSL#3™ combined with VDZ in patients with UC.

**Methods:** Clinical data were retrospectively collected from patients with active UC who received VSL#3™ combined with VDZ for at least 12 weeks at the Second Affiliated Hospital of Zhejiang University School of Medicine in China between January 2023 and December 2024. The primary endpoints were clinical response rates at weeks 6 and 12. Secondary endpoints included clinical remission, changes in inflammatory bowel disease questionnaire (IBDQ) scores, safety assessment.

**Results:** Using PRO2 criteria, clinical response rates were 82.4% (14/17) at 6 weeks and 100.0% (17/17) at week 12, with clinical remission in 58.8% (10/17) at week 12. By Full Mayo Score, clinical response and remission at week 12 were 68.8% (11/16) and 50.0% (8/16), respectively. These response rates appeared numerically higher than those reported in published historical VDZ monotherapy studies, although direct comparisons are limited by the single-arm design. Mean IBDQ score improved from 157.3 at baseline to 174.5 at week 12 (p<0.001). The combination was well-tolerated with no serious adverse events. Fatigue, borborygmus, and arthralgia were reported in 1/17 (5.9%), 2/17 (11.8%), and 1/17 (5.9%) patients, respectively. Notably, all 3 patients with baseline history of *Clostridioides difficile* (CDI) positivity tested negative for both toxin and antigen at 12 weeks; one toxin-positive patient had received anti-CDI antibiotic therapy.

**Conclusions:** This preliminary real-world study suggests that VSL#3™ may enhance early clinical outcomes in patients receiving VDZ for active UC. The observed benefits may be related to complementary effects on the gut microbiota and intestinal immune responses, although these mechanisms were not directly assessed. Further larger prospective randomized controlled trials are warranted to confirm these findings.

**Fund program:** Traditional Chinese Medicine Inheritance and Innovation Talent Support Program, Chinese Medicine Administration Bureau (2023ZR036); Youth Innovation Project of Zhejiang Provincial Health Department (2023RC168); Qingfeng Scientific Research Fund of the China Crohn’s & Colitis Foundation (CCCF-QF-2022C21-16).

## Introduction

Ulcerative colitis (UC) is a chronic relapsing inflammatory bowel disease characterized by abdominal pain, diarrhea, and mucous or bloody stools, leading to substantial impairment in quality of life ^[1]^. Gut microbiota dysbiosis and mucosal immune dysfunction are considered key contributors to the pathogenesis of UC ^[2]^. Vedolizumab (VDZ), a gut-selective biologic that blocks the interaction between α4β7 integrin and mucosal addressin cell adhesion molecule-1, thereby reducing intestinal inflammation while limiting systemic immunosuppressive effects ^[3]^.

Despite the established efficacy and favorable safety profile of VDZ, its relatively slow onset of action during induction therapy remains a clinical challenge, leaving some patients with a persistent symptomatic burden in the early treatment phase. Intestinal dysbiosis is increasingly recognized as a key contributor to the pathogenesis of ulcerative colitis. VSL#3™, a high-potency multi-strain probiotic formulation, has been reported to support epithelial barrier function, modulate mucosal immune responses, and promote restoration of gut microbial homeostasis^[4]^. However, whether adjunctive VSL#3™ could improve early outcomes during VDZ induction therapy remains unclear. Therefore, we conducted this pilot real-world study to evaluate the short-term efficacy and safety of combined VSL#3™ and VDZ therapy in patients with active UC and to describe exploratory *Clostridioides difficile* (CDI) -related observations.

## Study Subjects and Methods

### 1. Patients and Methods

This was a retrospective, single-arm, real-world preliminary study included patients with moderate active UC treated at the department of Gastroenterology, Second Affiliated Hospital of Zhejiang University School of Medicine, between January 2023 and December 2024, who completed 12 weeks of combined VDZ and VSL#3™ therapy.

Inclusion Criteria were as follows: (1) male or female aged 18-85 years; (2) diagnosis of UC according to the Consensus opinion on the Diagnosis and Treatment of inflammatory bowel disease (2018, Beijing); (3) diagnosis of UC at least 90 days before baseline, supported by comprehensive colonoscopy or sigmoidoscopy findings and histopathological findings obtained within the past year; available endoscopic and pathological reports within 3 months before baseline, and blood test results within 1 week before baseline; (4) active UC at screening, defined as a Mayo score ≥3; (5) completion of at least 12 weeks of probiotic VSL#3 ™ treatment by December 2024. Exclusion Criteria: (1) Diagnosis of Crohn’s disease, IBD-unclassified or other colitis; (2) Severe cardiac, hepatic, pulmonary, renal, hematologic, or other serious comorbidities that could affect study evaluation or patient safety, including malignancy, AIDS, significant renal dysfunction, urine protein >+, microscopic hematuria, alanine aminotransferase >2 × upper limit of normal, serum creatinine above the upper limit of normal, platelet count < 50*10^9^/L or white blood cell count < 3.0*10^9^/L; (3) Patients with uncontrolled systemic infections or severe enteric infections requiring immediate antibiotics treatment, dysplasia, or malignancy at baseline (but patients with recurrent CDI were eligible for enrollment if, in the investigators’ judgment, initiation of vedolizumab therapy remained clinically appropriate); (4) VSL#3™ treatment duration shorter than 12 weeks.

The study was approved by the Ethics Committee of the Second Affiliated Hospital of Zhejiang University School of Medicine (Approval No. YJ2024-0965).

### 2. Study Methods

#### 2.1 Data Collection

The Collected clinical data included sex, age, Montreal classification, laboratory parameters, endoscopic findings, CDI detection, concomitant and previous medication use, and adverse events.

#### 2.2 Intervention

All enrolled patients received VSL#3™ (450 billion CFU per sachet, Lot. No. G2I105) twice daily for 12 weeks, together with intravenous infusions of VDZ 300 mg (Takeda Pharmaceutical, Japan, registration number S20200006) at weeks 0, 2, and 6.

#### 2.3 Efficacy and Safety Evaluation

Clinical response and clinical remission were evaluated at weeks 6 and 12. Full Mayo Score and PRO2 scores were assessed at baseline before the first intervention (week 0) and after 12 weeks of combined VDZ and VSL#3™ treatment. The Full Mayo Score was used to assess disease activity in patients with UC. This four-item scale included stool frequency, rectal bleeding, endoscopic findings and the Physician’s Global Assessment, each scored 0-3, with higher scores indicating more severe disease. Endoscopic remission was defined as a Mayo endoscopic subscore of 0 or 1, and complete endoscopic healing was defined as a Mayo endoscopic subscore of 0. The doctor assesses the patient’s condition: Based on the patient’s disease situation, a score of 0 indicates normal, 1 indicates mild, 2 indicates moderate, and 3 indicates severe. This scale corresponds to the four-item Mayo scoring system used in the Chinese National Clinical Guideline for Ulcerative Colitis.

Clinical Response according to PRO2 criteria was defined as a ≥50% reduction in the PRO2 score, based on rectal bleeding and stool frequency. Clinical response according to the Full Mayo Score was defined as a ≥30% and ≥3-point reduction from baseline, accompanied by a ≥1-point decrease in the rectal bleeding subscore or an absolute rectal bleeding subscore of 0 or 1. Clinical Remission as according to PRO2 criteria was defined as rectal bleeding and stool frequency subscores of 0. Clinical remission according to the Full Mayo Score was defined as a total score ≤2, with no individual subscore >1. Safety was assessed by monitoring adverse events during treatment, including infusion reactions, infections, hepatic or renal impairment, nausea, and other symptoms. Infusion reactions were defined as adverse events occurring on the day of VDZ infusion or on the following day.

Non-serious AEs were defined as medical events that did not lead to discontinuation of VDZ or VSL#3™, or hospitalization. Serious AEs were defined as those resulting in hospitalization or patient death.

### 3. Statistical Analysis

Data were analyzed using SPSS version 20.0. Normally distributed continuous variables were presented as mean ± standard deviation (SD), whereas non-normally distributed continuous variables were presented as median and interquartile range (IQR). Paired comparisons between baseline and week 12 were performed using the Wilcoxon signed-rank test for non-normally distributed continuous variables. Categorical variables were presented as frequencies and percentages. The chi-square test or Fisher’s exact test was used when appropriate. Analyses were performed based on the available evaluable data for each endpoint. A p-value < 0.05 was considered statistically significant.

## Results

### Patients and characteristics

Between January 2023 and December 2024, 18 patients with moderate active UC were enrolled at a single tertiary hospital in China. One patient withdrew because of disease progression, and 17 patients were included in the final analysis. The final analysis population had a mean age of 39.2 ±16.7 years and was predominantly male (82.4%, 14/17). The mean baseline Full Mayo Score was 6.7 ±1.2. Most patients had extensive colitis (E3, 11/17, 64.7%), while 6/17 (35.3%) had left-sided colitis (E2). No patient had proctitis (E1). Among them, 15 patients had initial-onset disease and 2 patients had chronic relapsing type. Previous medications included 5-aminosalicylic acid (5-ASA) in all 17 patients (100%), corticosteroids in 1 patient (5.9%), and traditional Chinese medicine in 1 patient (5.9%). No patients had previously used immunosuppressants or biologics. In addition, 14/17 patients continued oral mesalamine without dose adjustment during the treatment period (Table 1).

**Table 1.**
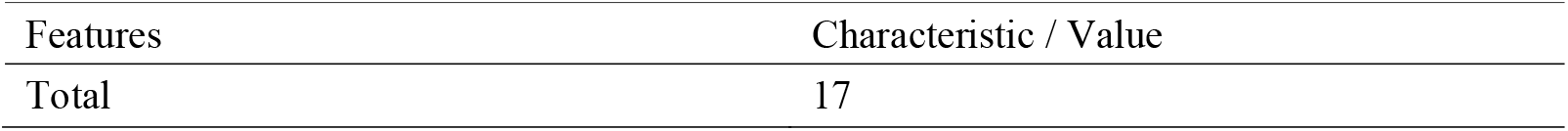
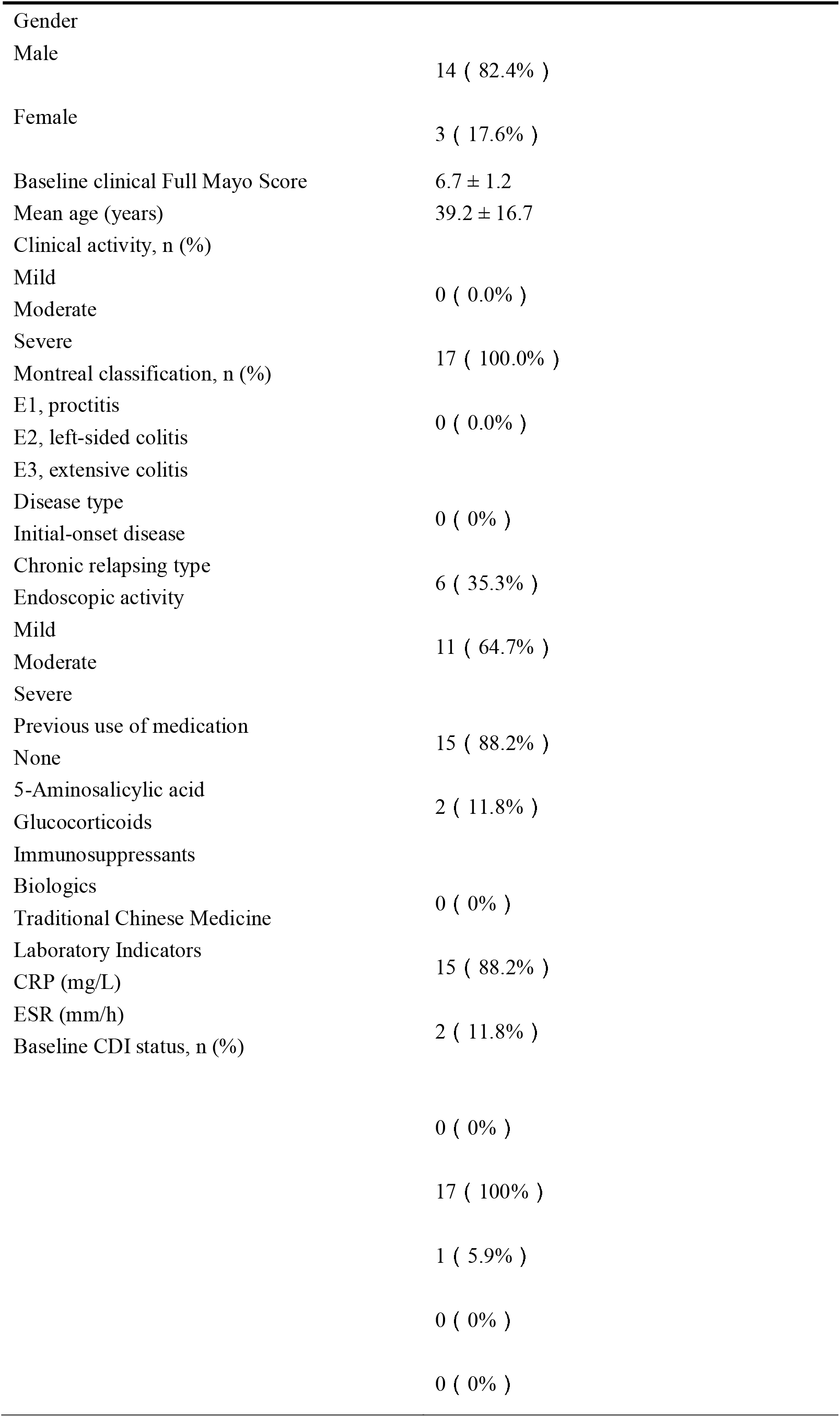
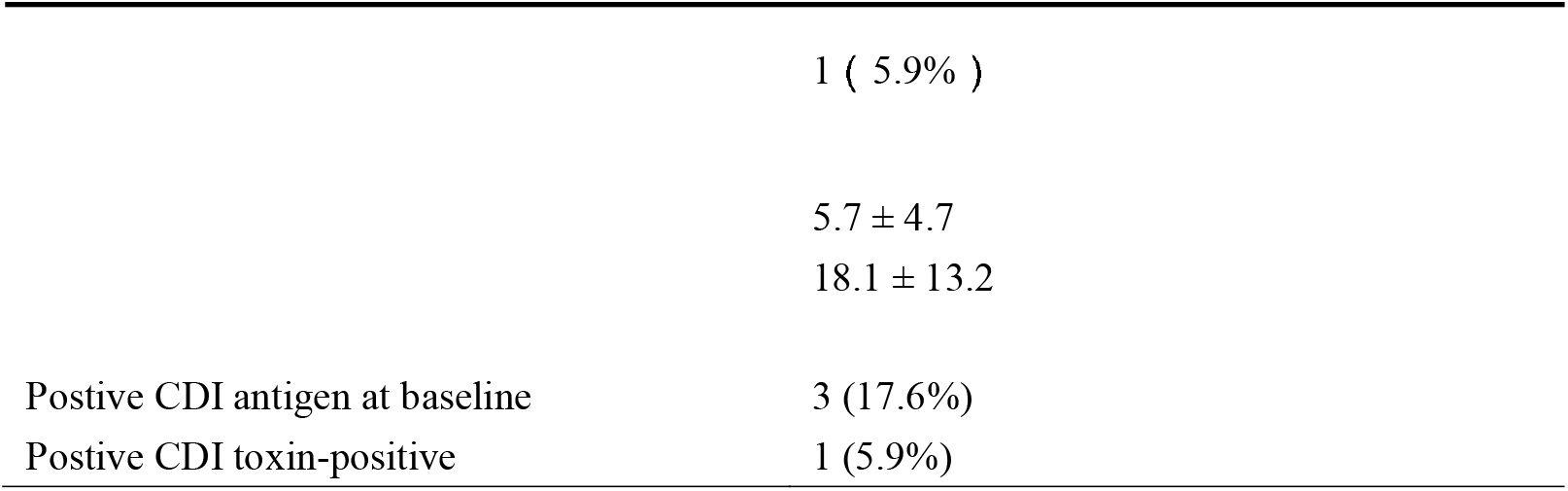
Baseline demographic and clinical characteristics of patients with UC.

| Features | Characteristic / Value |
| --- | --- |
| Total | 17 |
| Gender |  |
| Male | 14 ( 82.4% ) |
| Female | 3 ( 17.6% ) |
| Baseline clinical Full Mayo Score | 6.7 ± 1.2 |
| Mean age (years) | 39.2 ± 16.7 |
| Clinical activity, n (%) |  |
| Mild | 0 ( 0.0% ) |
| Moderate |  |
| Severe | 17 ( 100.0% ) |
| Montreal classification, n (%) |  |
| E1, proctitis | 0 ( 0.0% ) |
| E2, left-sided colitis |  |
| E3, extensive colitis |  |
| Disease type | 0 ( 0% ) |
| Initial-onset disease |  |
| Chronic relapsing type | 6 ( 35.3% ) |
| Endoscopic activity |  |
| Mild | 11 ( 64.7% ) |
| Moderate |  |
| Severe |  |
| Previous use of medication | 15 ( 88.2% ) |
| None |  |
| 5-Aminosalicylic acid | 2 ( 11.8% ) |
| Glucocorticoids |  |
| Immunosuppressants |  |
| Biologics | 0 ( 0% ) |
| Traditional Chinese Medicine |  |
| Laboratory Indicators | 15 ( 88.2% ) |
| CRP (mg/L) |  |
| ESR (mm/h) | 2 ( 11.8% ) |
| Baseline CDI status, n (%) |  |
|  | 0 ( 0% ) |
|  | 17 ( 100% ) |
|  | 1 ( 5.9% ) |
|  | 0 ( 0% ) |
|  | 0 ( 0% ) |
|  | 1 ( 5.9% ) |
|  | 5.7 ± 4.7 |
|  | 18.1 ± 13.2 |
| Postive CDI antigen at baseline | 3 (17.6%) |
| Postive CDI toxin-positive | 1 (5.9%) |

Baseline laboratory parameters showed a mean C-reactive protein (CRP) level of 5.7 ± 4.7 mg/L and mean erythrocyte sedimentation rate (ESR) of 18.1 ± 13.2 mm/h. Three patients tested positive for CDI at baseline, and they were clinically stable and did not require hospitalization or escalation of antibiotics during enrollment. One patient was toxin-positive and received a 2-week course of anti-CDI antibiotic therapy, whereas two patients were antigen-positive but toxin-negative and did not receive antibiotic treatment. By Week 12, all three patients tested negative for both CDI toxin and antigen (Table 2).

**Table 2.** Clinical characteristics and follow-up CDI testing results of the three patients with baseline CDI positivity.

| Case | Baseline CDI test | Symptoms | Anti-CDI antibiotics | Week 12 CDI test | Full Mayo Score change |
| --- | --- | --- | --- | --- | --- |
| 1 | Antigen positive, toxin negative | Loose stools with a little blood 3-4 times/day | No | Antigen and toxin negative | - 4 |
| 2 | Antigen positive, | Loose stools with | No | Antigen and toxin | - 6 |
|  | toxin<br>negative | a little<br>blood 3-4<br>times/day |  | negative |  |
| 3 | Antigen and<br>toxin<br>positive | Diarrhea<br>with bloody<br>stools 5-7<br>times/day | Yes | Antigen and<br>toxin<br>negative | - 5 |

### 2. Efficacy analysis

Figure 1A shows the clinical response and remission rates evaluated using PRO2 criteria at weeks 6 and 12. Using PRO2 criteria, 14/17 patients (82.4%) achieved clinical response at week 6, increasing to 17/17 patients (100.0%) at week 12. According to the Full Mayo Score at week 12 11/16 patients (68.8%) achieved clinical response, and 8/16 patients (50.0%) achieved clinical remission. Endoscopic remission was also observed in 8/16 patients (50.0%), as assessed using the Mayo endoscopic subscore. One patient completed the treatment period but declined the week 12 hospital reassessment; therefore, Mayo score-based outcomes were evaluated in 16 patients.

**Fig 1.**
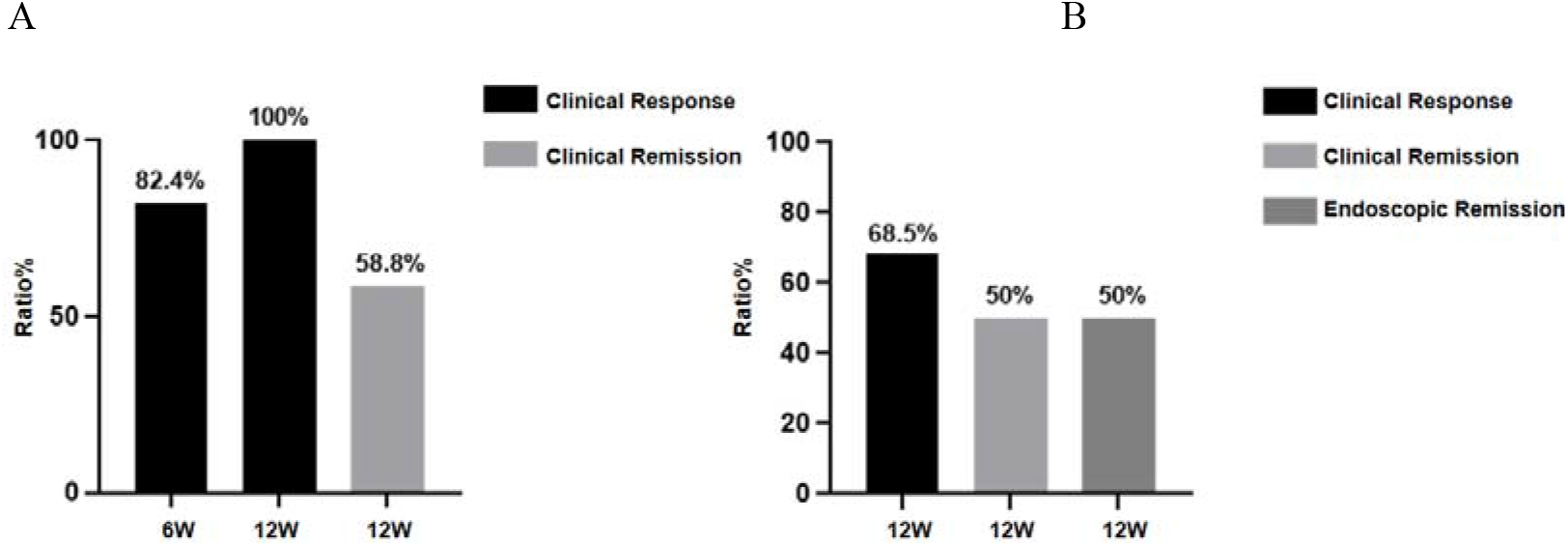
Clinical response and remission rates. (A) Clinical response and remission rates evaluated using PRO2 criteria at weeks 6 and 12. (B) Clinical response and clinical remission evaluated using the Full Mayo Score, and endoscopic remission evaluated using the Mayo endoscopic subscore at week 12.

These results suggest that most patients experienced symptomatic improvement after combination therapy. Changes in Full Mayo Score and IBDQ scores from baseline to week 12 are shown in Figures 2A and 2B.

**Fig 2.**
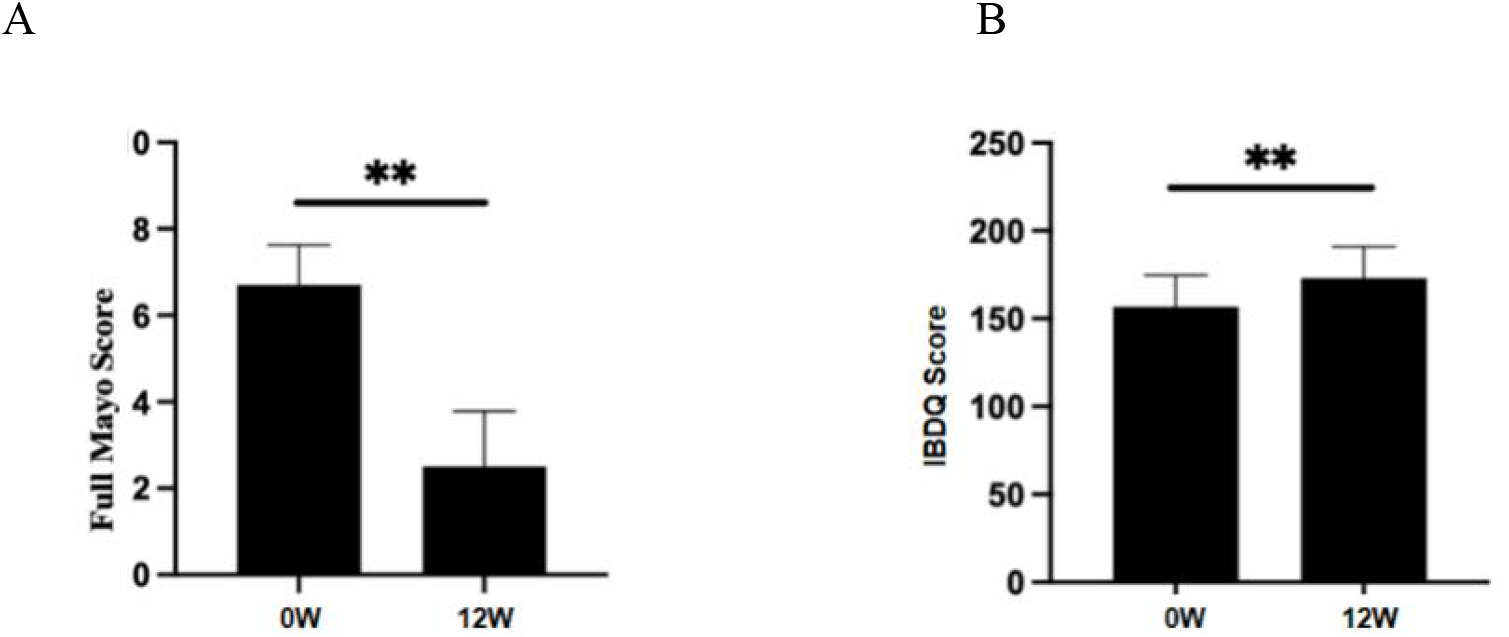
Changes in clinical and quality-of-life scores from baseline to week 12. (A) Full Mayo Score. (B) Inflammatory Bowel Disease Questionnaire (IBDQ) scores.

Among the 16 patients with evaluable Full Mayo Score, the mean Full Mayo Score decreased from 6.7 ± 1.2 at baseline to 2.6 at week 12, showing a significant reduction (P < 0.001). The mean IBDQ score increased from 157.3 at baseline to 174.5 points at week 12, indicating improved quality of life (P < 0.001).

#### Safety Assessment

A total of 18 patients were included in the study. One patient experienced disease progression during treatment and was switched to infliximab, leading to study withdrawal. Safety was therefore assessed in the remaining 17 patients who completed 12 weeks of treatment. Overall, the combination therapy was generally well tolerated. During the treatment period, fatigue, borborygmus, and arthralgia were reported in 1/17 (5.9%), 2/17 (11.8%), and 1/17 (5.9%) patients, respectively. All reported symptoms resolved spontaneously or after symptomatic treatment. No serious adverse events were observed. All three patients with baseline CDI positivity tested negative for both CDI antigen and toxin at week 12; one toxin-positive patient had received anti-CDI antibiotic therapy.

#### Discussion

In this retrospective real-world study, combined VSL#3™ and VDZ therapy was associated with early clinical improvement in patients with active UC. Using PRO2 criteria, clinical response rates were 82.4% at week 6 and 100% at week 12, with a clinical remission rate of 58.8% at week 12. When assessed using the Full Mayo Score, clinical response and remission rates at week 12 were 68.8% and 50.0%, respectively.

Our group previously reported the long-term clinical response and remission of VDZ monotherapy in Chinese patients with UC at week 54 was 88.1% (74/84), 59.5% (47/84), respectively^[4]^. The present study extends these observations by exploring whether adjunctive probiotic therapy may improve early outcomes during VDZ induction. To our knowledge, this is one of the first real-world studies to evaluate the combination of VSL#3™ and VDZ in patients with moderate UC. The clinical response rates observed in our study appeared numerically higher than those reported in previous studies of VDZ monotherapy; however, this comparison should be interpreted cautiously because of differences in study design, patient populations, and response definitions. In a randomized double-blind placebo-controlled trial of VDZ in patients with moderate-to-severe UC, clinical response and remission rates at week 6 were 47.1% and 16.9%, respectively ^[5]^. A study evaluating VDZ using patient-reported outcomes (PRO2) reported a clinical response rate of 73.4% (47/64) at week 14 ^[6]^. In a head-to-head trial comparing VDZ with adalimumab, the VDZ group achieved a clinical response rate of 67.1% (257/383) and a clinical remission rate of 26.6% at week 14 ^[7]^. In our study, symptom-based clinical response rates at weeks 6 and 12 were numerically higher than those reported in these previous VDZ studies, although direct comparisons are limited by differences in design, baseline characteristics, and endpoint definitions.

Gut microbiota disruption is considered an important contributor to UC pathogenesis. The clinical effects of probiotics in UC may vary according to strain composition, dose, and formulation. A recent meta-analysis suggested that selected multistrain probiotics containing Lactobacillus and Bifidobacterium may help induce endoscopic remission and prevent clinical relapse in UC^[8]^.

VSL#3™ is a high-concentration multi-strain probiotic mix containing one strain of *Streptococcus thermophilus* BT01, three strains of Bifidobacteria (*B. breve* BB02; *B. animalis* subspecies *lactis* BL03; and *B. animalis* subspecies *lactis* BI04), and four strains of Lactobacilli (*L. acidophilus* BA05, *L. plantarum* BP06, *L. paracasei* BP07, and *L. helveticus* BD08)^[9]^. The potential mechanisms may involve synergistic effects of the eight bacterial strains on tight junction proteins, intestinal epithelial barrier function, and mucosal immune-inflammatory pathways ^[10]^. Therefore, we hypothesize that combining VSL#3™ with VDZ may enhance the interaction between beneficial bacteria and T cells, providing a dual effect: rapid restoration of the gut microbiota and regulation of the immune system. This combination may contribute to improvements in frequency and rectal bleeding, support clinical remission, and potentially improve quality of life. However, microbiota composition, epithelial barrier markers, and immune cell profiles were not directly assessed in the present study; thus, mechanistic interpretations should be considered exploratory.

Previous studies have evaluated the risk of gastrointestinal infections, including CDI, in UC patients treated with VDZ ^[11]^. This may be clinically relevant given the role of gut-selective T-cell trafficking inhibition in intestinal immune surveillance ^[12].^ Concomitant VSL#3™ treatment may be relevant in this context because of its potential effects on gut microbiota and CDI-related pathways. Transcriptomic data have suggested a potential protective role of lactobacilli against CDI, by reducing virulence-related pathways, including motility, quorum sensing, survival and spore germination potential ^[12]^. In our cohort, three patients were CDI-positive at baseline: one was toxin-positive and received anti-CDI antibiotic therapy, whereas two were antigen-positive but toxin-negative and did not receive antibiotics. All three tested negative for both CDI antigen and toxin at week 12. Given the very small number of cases, the absence of a control group, and the use of antibiotic therapy in one patient, no conclusion can be drawn regarding the effect of VSL#3™ on CDI status. However, this exploratory observation may warrant further investigation. This study has several limitations. First, its retrospective single-center design and small sample size limit the generalizability of the findings. Second, the absence of a contemporaneous VDZ monotherapy control group precludes definitive conclusions regarding the additive efficacy of VSL#3™. Third, microbiome analyses were not performed, preventing mechanistic exploration of probiotic-related effects. Finally, CDI-related findings were exploratory and based on a very limited number of cases, and therefore should be interpreted with caution.

In conclusion, combined VSL#3™ and VDZ therapy was associated with rapid symptomatic improvement and favorable short-term clinical response and remission rates in patients with moderate active UC. These preliminary real-world findings suggest that this combination may represent a potential therapeutic strategy, but prospective controlled studies are required to confirm its efficacy, safety, and long-term benefits.

## Data Availability

The data supporting the findings of this study are not publicly available because they contain sensitive patient information and their disclosure could compromise patient privacy. De-identified data may be obtained from the corresponding author upon reasonable request, subject to approval by the relevant ethics committee and institution and compliance with applicable data protection requirements.

## Author Contributions

*Jiakai Luo*: Writing of the manuscript, manuscript revision, case collection.

Qiao Yu, Xiaoying Wang, Jiakai Luo, Dingting Xu, Hanyun Zhang, Miaoyan Chen, Shuyan Li: Data organization, statistical analysis.

Qiao Yu, Yan Chen: Manuscript revision.

Parnia Ghanad, Maryam Maleki Goli: Writing – review & editing

